# Factors Associated with Outcomes of Inpatient Severe Malaria Cases in the Ashanti Region, Ghana: An Analytic Cross-sectional Study using Routine Surveillance Data, 2018 to 2022

**DOI:** 10.64898/2026.03.26.26349387

**Authors:** Courage Eli Yevugah, Michael Opoku-Mireku, Bismark Sarfo, Harriet Affran Bonful

## Abstract

**Background:** Malaria remains a major global health threat, with 249 million cases and 609,000 deaths reported in 2022. The Ashanti Region of Ghana bears a disproportionate burden, with severe malaria accounting for 24% of hospital admissions in 2021, above the national average of 19%. Despite intensified control efforts, inpatient mortality patterns remain poorly understood. This study identifies key determinants of severe malaria mortality among hospitalized patients in the Ashanti Region.

**Methods:** We analyzed inpatient surveillance data from the District Health Information Management System 2 (DHIMS2) for severe malaria admissions from 2018 to 2022. Descriptive statistics, bivariate analyses with robust survey estimation (accounting for design effects), and multivariable Firth penalized logistic regression were used to identify mortality predictors. Survey-adjusted logistic regression served as a sensitivity analysis to validate findings.

**Results:** Among 54,544 severe malaria admissions, females comprised 51.1% and children under five 39.4%. The case fatality rate was 0.4% (200 deaths). Mortality was significantly associated with age, occupation, insurance status, facility ownership, admitting department, length of stay, and comorbidities. Males had 1.4 times higher mortality odds than females. Compared to children under five, patients aged 5–17 years had 44% lower odds of mortality (aOR = 0.56, 95% CI: 0.33–0.94). Active NHIS membership had lower mortality odds by 67% (aOR=0.33, 95% CI: 0.25–0.45) compared to inactive membership. Admissions to faith-based facilities showed lower mortality odds (aOR=0.38, 95% CI: 0.23–0.65) than government facilities, while medical wards had higher odds (aOR=2.38, 95% CI: 1.48–3.84) than paediatric wards. Stays of 3–5 days were associated with lower mortality odds (aOR=0.67, 95% CI: 0.47–0.97) compared to stays <3 days. Those with comorbidities had twice the mortality odds versus those without. Sensitivity analysis confirmed consistent direction and significance.

**Conclusion:** Age, comorbidities, insurance coverage, facility type, and admission practices strongly influence severe malaria mortality in Ashanti. Strengthening NHIS enrollment, extending inpatient monitoring beyond three days, and adopting best practices from paediatric and faith-based facilities could improve survival. Integrating comorbidity screening and management into malaria protocols is essential to reducing preventable deaths.

## Introduction

Malaria remains a critical global health challenge, particularly in sub-Saharan Africa, where it causes substantial morbidity and mortality (1,2). Approximately 3.2 billion people, half the world’s population, are at risk (3). In 2021, an estimated 247 million cases and 619,000 deaths occurred worldwide, with sub-Saharan Africa accounting for over 90% of the burden (4).

Severe malaria is a medical emergency requiring prompt diagnosis and treatment to prevent death and disability (White, 2022). It carries a high case fatality rate (CFR), approximately 90% outside health facilities and 20% in facilities (6), with many deaths occurring before patients reach care (7). Severe cases often require admission due to complications such as respiratory distress, cerebral malaria, acute kidney injury, anaemia, bleeding disorders, and co-infections (8). Before the 2019 malaria vaccine rollout, incidence among infants and young children in sub-Saharan Africa was estimated at 1,919 per 100,000 person-years, with attributable mortality of 231 per 100,000 per year (9). In Ghana, malaria remains a leading cause of morbidity and mortality, with approximately 5 million cases reported in 2022 (4), accounting for 30% of outpatient visits and 23% of inpatient admissions (10). In 2020, an estimated 308,887 severe malaria cases and 308 deaths were recorded (11). A quarter of malaria deaths among children occur in those under five (12). Despite declines, Ghana fell off track in meeting Global Technical Strategy milestones (4).

DHIMS2 data, Ghana’s primary malaria surveillance system (1,13)show the Ashanti Region consistently experiences higher severe malaria incidence than the national average. In 2021, it recorded 24% (98,148) of severe malaria admissions compared to the national 19% (24,508), and 42 malaria-related deaths versus the national average of 20. In 2020, inpatient admissions were 20% (63,991) in Ashanti compared to 18% (19,306) nationally. This pattern persisted despite intensified control measures, including ITN distribution, IRS, IPT, case management, and surveillance (14–17). This reversal in progress is largely attributed to COVID-19 disruptions to malaria services, affecting supply chains, healthcare access, and commodity availability (18). Other factors include humanitarian emergencies, insufficient investment, and socioeconomic, demographic, and geographic inequalities, all exacerbated by the pandemic (19,20).

Understanding inpatient severe malaria characteristics and outcomes is vital for targeted interventions and resource allocation in Ashanti and Ghana. Previous studies highlight factors such as age, education, health insurance, comorbidities, and facility ownership influencing outcomes (21–24). However, context-specific factors in Ashanti remain underexplored.

This analytic cross-sectional study uses DHIMS2 inpatient records from 2018 to 2022 to assess severe malaria outcomes among inpatients in Ashanti. No prior study examines inpatient severe malaria characteristics and outcomes in this region using DHIMS2 data. It is expected that critical evidence will inform further studies to improve sustainable malaria control in Ghana.

This study aims to describe the characteristics of inpatients with severe malaria and identify factors associated with admission outcomes in the Ashanti Region from 2018 to 2022.

## Materials and Methods

### Study Design

This study employed an analytic cross-sectional design using secondary inpatient data on severe malaria from DHIMS2 for the Ashanti Region between 2018 and 2022.

### Study Settings

This study was conducted in the Ashanti Region of Ghana, which stretches over 24,389 square kilometres, comprising 10.2% of Ghana’s land area. With an estimated population of 5,440,463 (17.6%), it is the second most populous region in Ghana (30,832,019), after the Greater Accra region (17.7%) as per Ghana’s 2021 Population and Housing census (25).

The region experiences two distinct climatic variations, with an annual rainfall range of 1,200 to 1,700mm, between April and November, with peaks in July and October. The dry season, influenced by the harmattan winds, extends from December to March. Temperature ranges from an average of 27°C in the southern districts to 32°C in the northern districts, peaking in February. These climatic conditions significantly influence malaria transmission dynamics in the region.

The region features a mix of moist semi-deciduous forest and guinea savannah, creating a transitional zone. The southern areas have dense forests, while the north is more savannah-like. Environmental changes are driven by settlement expansion, artificial water bodies, and bushfires. This area contains 69% of Ghana’s forest reserves, including Tano Offin and Bobiri Forest reserves. Malaria prevalence among children under five is 8% (26) and case fatality in the region is 0.02% (27), as of 2022.

DHIMS 2 is a web-based tool implemented by the Ghana Health Service (GHS) based on the District Health Information System 2 (DHIS 2) application software. It is used to capture, collate and analyze health-related data and disseminate reports over time across health institutions using tables, graphs, maps and other visualization methods. The DHIMS 2 is managed by the Center for Health Information Management, Ghana (CHIM-GH).

**Figure 1.**
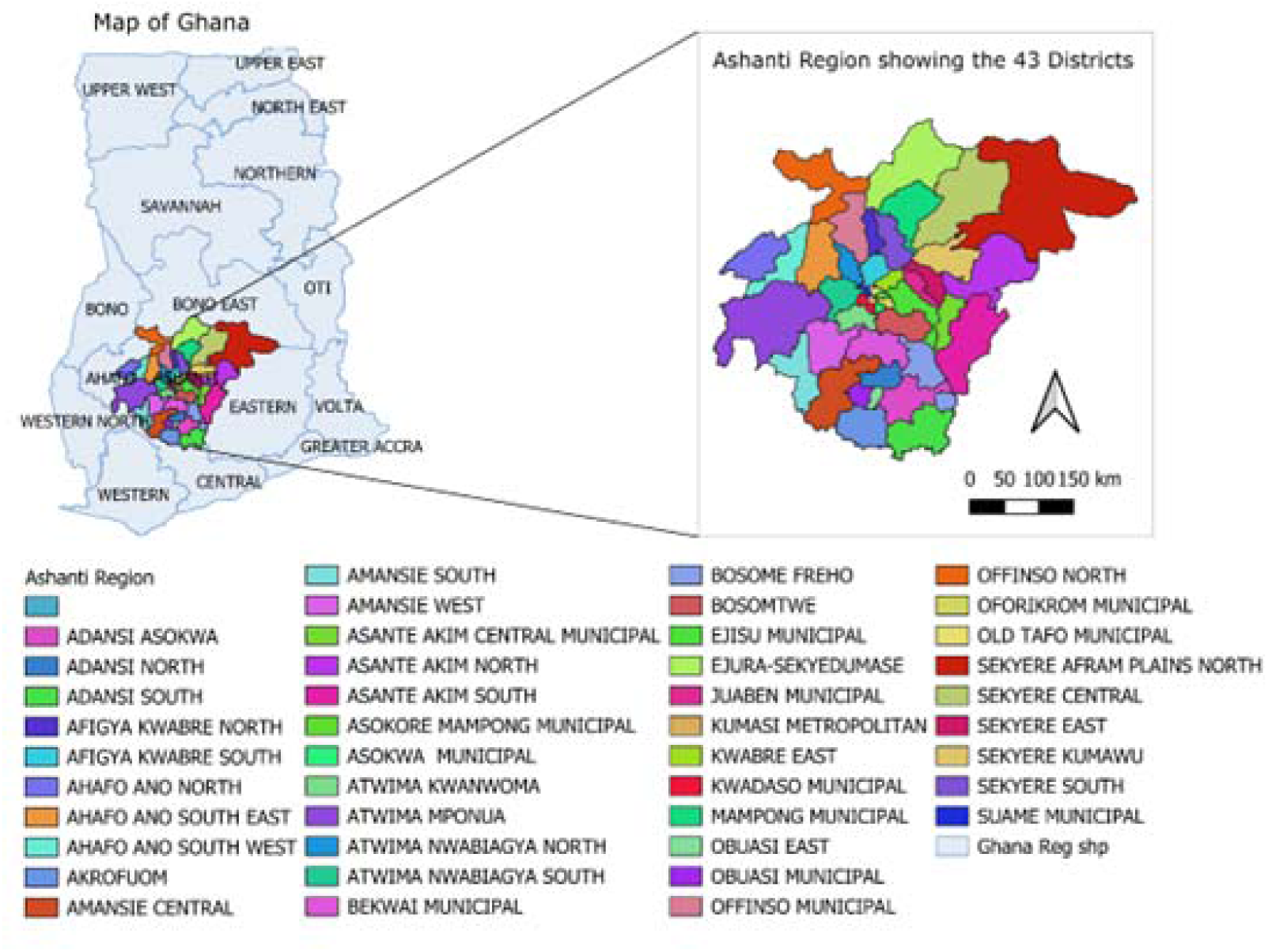
Map of Ghana showing Ashanti region and the 43 districts in this study. The Ashanti Region consists of 43 Metropolitan, Municipal, and District Assemblies (MMDAs), each led by a Chief Executive and a Member of Parliament (Figure 1). It has 1,654 health facilities, including CHPS compounds, clinics, health centres, district hospitals, and tertiary hospitals. The doctor-to-population ratio is 1:7,500, and the nurse-to-population ratio is 1:450. Health data is collected through the District Health Information Management System 2 (DHIMS 2).

### Study Population

This study included records of patients of all ages diagnosed with severe malaria and admitted to health facilities in the Ashanti Region from January 1, 2018, to December 31, 2022. Severe malaria is diagnosed by trained prescribers based on the Ghana Standard Treatment Guidelines, which are aligned with WHO criteria.

Malaria diagnosis is made by parasite confirmation using the blood smear microscopy with the Giemsa staining method and malaria RDT kits (28,29). There are different kinds of RDT kits used, but the Histidine-rich protein 2 (HRP2) is the most commonly used in health facilities in Ghana (29–32). Health facilities below hospital status provide pre-referral management and transfer patients to the hospital for admission and treatment. Admission records are entered into the inpatient registers and subsequently reported in DHIMS 2 after validation by facility validation teams or management.

### Inclusion and Exclusion Criteria

All inpatient records of severe malaria cases from January 1, 2018, to December 31, 2022, were included in the analysis. Records with missing admission outcomes (blanks), unspecified outcomes, absconded clients, and transferred out (referred) for admission outcome were excluded.

### Sample Size and Sampling

The largest required minimum sample size was computed using Cochran’s formula (33), yielding an estimated 385 cases. However, a census approach was adopted to enhance statistical power, including all eligible severe malaria records within the study period.

### Data Sources and Management

Data were extracted from the DHIMS 2 through the GHS Central Health Information Management (CHIM) unit, using structured extraction tools. The dependent variable was severe malaria admission outcome, categorized as “died” or “did not die”. Independent variables included demographic factors (sex and age) socio-economic factors (education, occupation, NHIS status, and residence), health facility characteristics (ownership, level of care, department), and clinical variables (length of stay, comorbidities, and district setting).

Initial data cleaning was conducted in Excel 365 (Microsoft Corporation), where data were checked for completeness, duplication, accuracy, and consistency. Anomalies were flagged for review and addressed accordingly. Length of stay was derived from admission and discharge dates, and districts were classified as urban or rural based on the criterion that more than 50% of the population resided in a given category using the population data in the 2021 Population and Housing Census report (25). Comorbidities were grouped based on the NHIA tariff classifications, which use the Ghana Diagnostic-Related Groupings (G-DRG) categories (34,35). Further management was conducted in Stata Version 17/BE (Stata Corporation LLC, Texas, USA, 2021), where categorical variables, including sex, educational status, etc., were encoded. Age was categorized into predefined groups: <5, 5-17, 18 years and above, and ‘Length of Stay’ was categorized into three groups: less than 3 days, 3-5 days, and greater than 5 days to facilitate succinct data handling and clear analysis. Variables were categorized based on established classifications and health system frameworks, and the literature (34,36,37).

### Statistical Analysis

Data analysis was executed using Stata. A robust estimation technique was employed, given the clustering of patients within health facilities, to account for intra-class correlation effects. This adjustment was vital to accurately estimating the standard errors and overall significance of the results. Using the survey data analysis in Stata, the data were declared as survey data with the “svyset” command, specifying the facility identifier as the primary sampling unit and the facility level as the strata. Descriptive and bivariate analyses were performed using the “svy” command.

Descriptive statistics were performed using frequencies and percentages. Missing values within variables were reported. Bivariate analysis was performed using the Chi-square test to discern the relationship between the outcome variable and independent variables. Univariable and multivariable logistic regression analyses were performed to determine the strength and direction of factors associated with severe malaria deaths. Variables demonstrating a p-value of less than 0.15 after assessing all independent variables in the univariable analyses, and theoretically important variables were included in the multivariable model using Firth’s penalized likelihood logistic regression, given the rare outcome (< 5%) prevalence in Ghana (38). This approach accounts for bias that may arise due to rare outcomes and separation and provides stable parameter estimates (39,40). Survey-adjusted logistic regression analyses “svy: logistic” were also performed as a sensitivity analysis to account for clustering and complex survey design. Full results of these analyses are presented in Additional file 1. The level of significance was set at an alpha of 0.05.

### Results

A record of 55,379 severe malaria cases was obtained from the DHIMS 2 inpatient event register through CHIM of GHS using the data extraction tool. 139 of the records were excluded because they were outside the inclusion dates of the study period. Out of the remaining 55,240 cases, 696 were excluded, for which the final outcome could not be ascertained. These included 521 transferred cases, 92 absconded cases, and 83 unspecified cases (Figure 2). The cases were reported from 65 health facilities in 50 different locations from 31 MMDAs of the Ashanti region.

**Figure 2.**
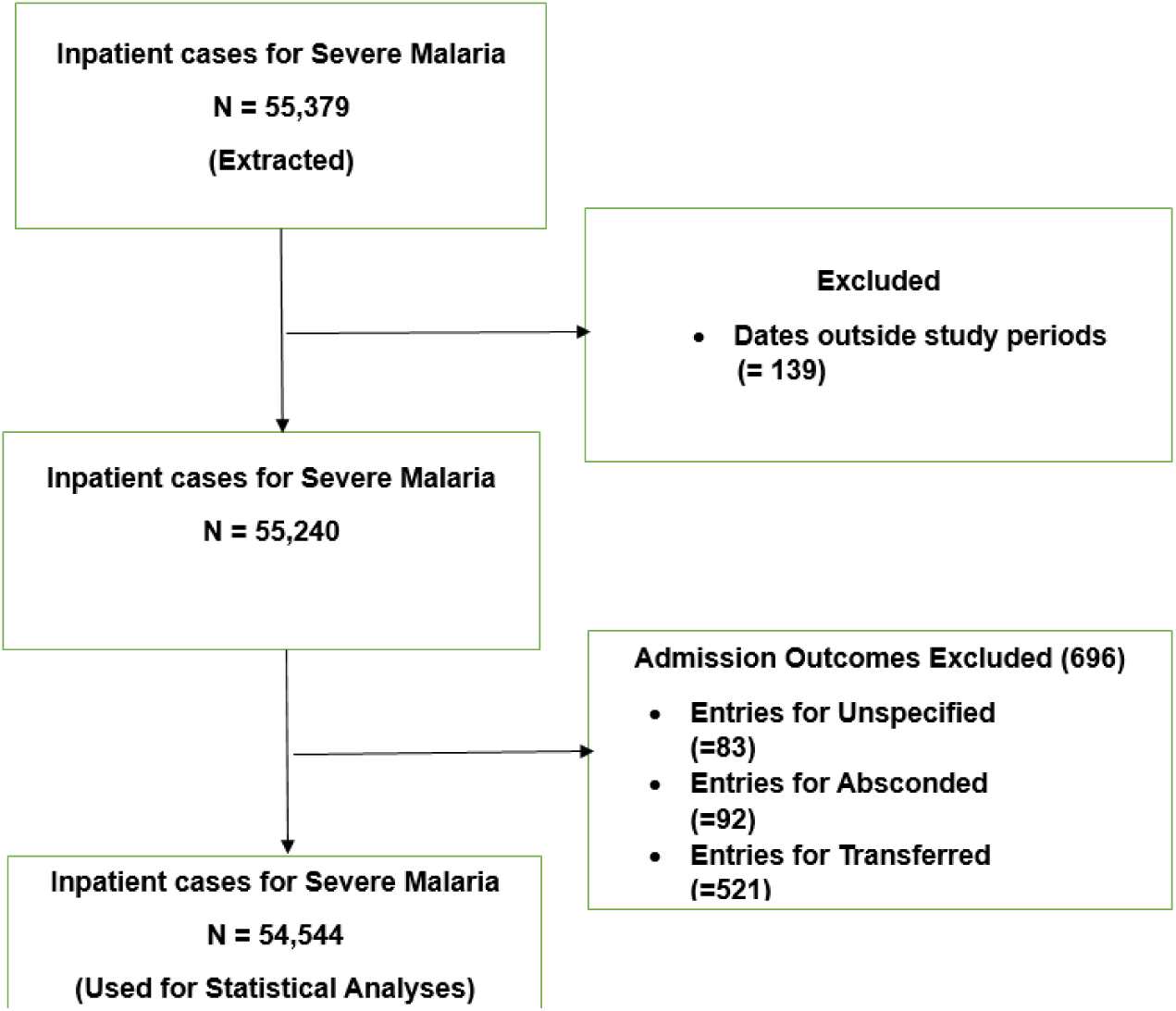
Study flowchart of severe malaria inpatient cases in the Ashanti region, 2018-2022. In total, 54,544 patients were included in the study, and more than half (51.1%) were females. Most patients were under the age of 5 years (39.4%), followed by the 5-17 age group (31.4%). A substantial proportion of the patients had no formal education (54.9%). Most patients (94.1%) were engaged in one occupation or another, among which students (72.8%) were the majority, and less than five per cent (3.2%) were unemployed. Most patients were active members of the NHIS (85.4%). Most facilities were government-owned (81.4%) and primary-level (98.7%). Regarding length of stay, 69.3% of the patients stayed for less than 3 days. The department in the facilities to most frequently admit severe malaria cases was the paediatric department (48.7%), followed by the medical department (47.7%). Among the patients, 42.5% presented with comorbidities, which included anaemic disorders (12.3%), septicaemia/systemic infections (7.1%), lower respiratory disorders (5.7%), diarrhoea and vomiting disorders (4.6%), among others. More than half of the patients were from urban districts (52.1%) (Table 1).

**Table 1.**
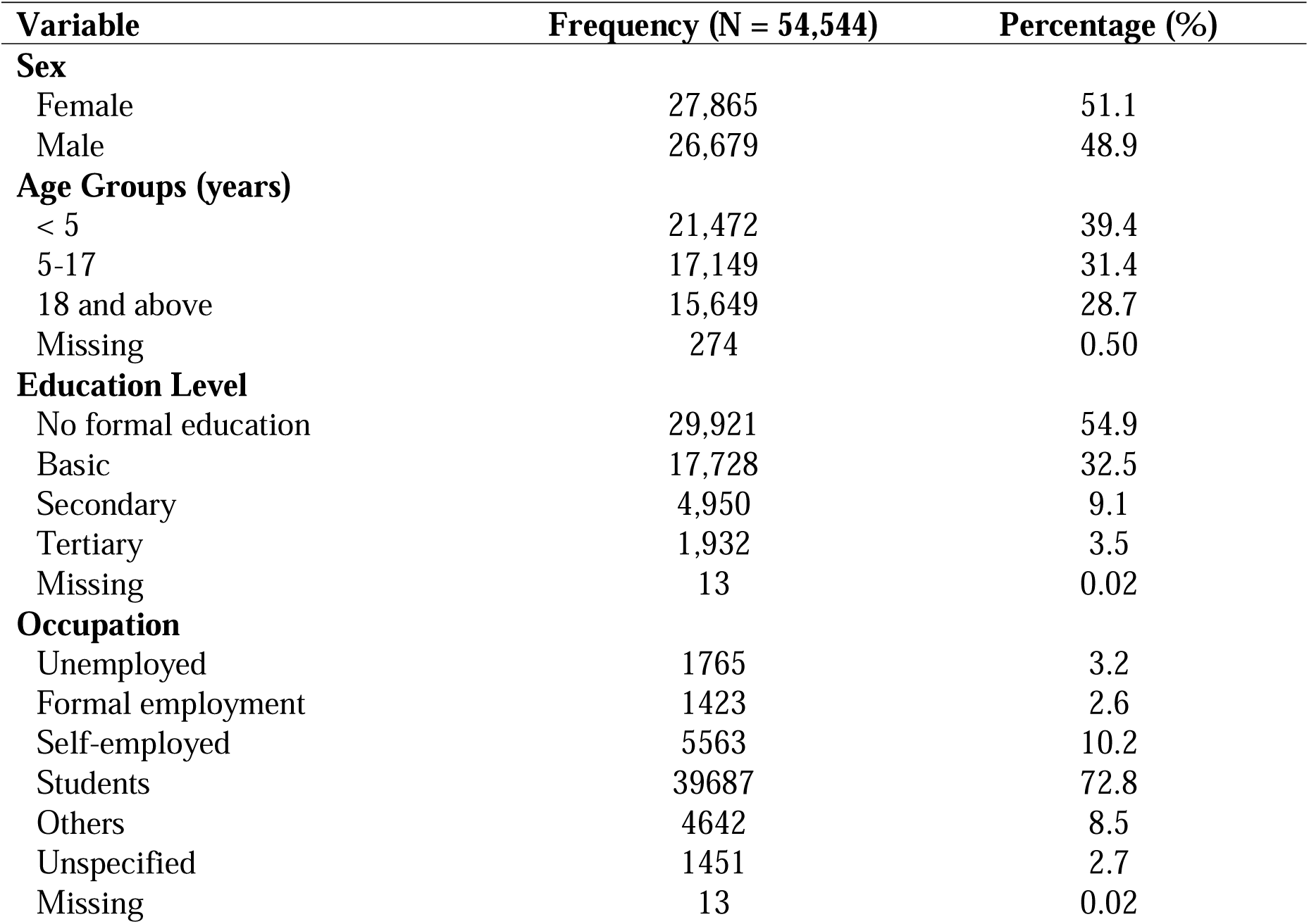
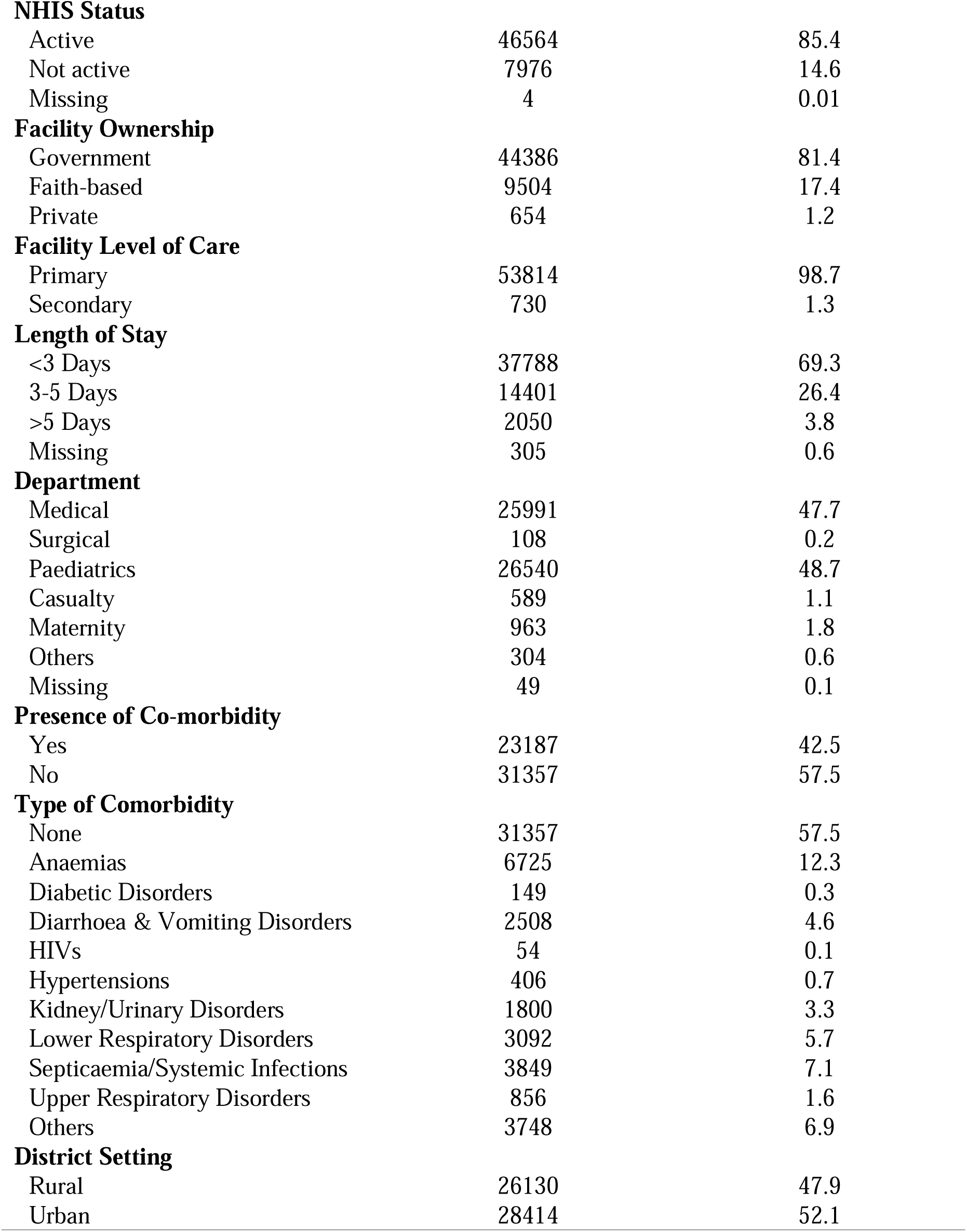
Characteristics of Inpatient severe malaria in the Ashanti region, 2018 – 2022.

Characteristics and bivariate analysis of severe malaria outcomes showed a 0.4% case fatality rate (200 deaths). Males comprised 52.5% of deaths, though not statistically significant (L2 = 0.76, p 0.387). Age distribution was significantly associated with outcomes (L2 = 52.76, p < 0.001), with the highest deaths in patients above 18 years (63.0%). Facility care level (L2 =17.4, p < 0.001) and stay duration (L2 = 3.21, p = 0.047) were significantly associated with outcomes, with 75.9% of deaths occurring within three days. Comorbidities were significantly associated with outcomes (L2 = 17.91, p < 0.001), with 60.0% of deceased patients having comorbidities, mainly anaemias (13.5%). Deaths occurred mostly in medical (73.9%) and paediatric departments (19.1%), though not statistically significant (L2 = 2.8, p = 0.058). Educational level, facility ownership, and district settings showed no significant differences in outcomes (Table 2).

**Table 2.**
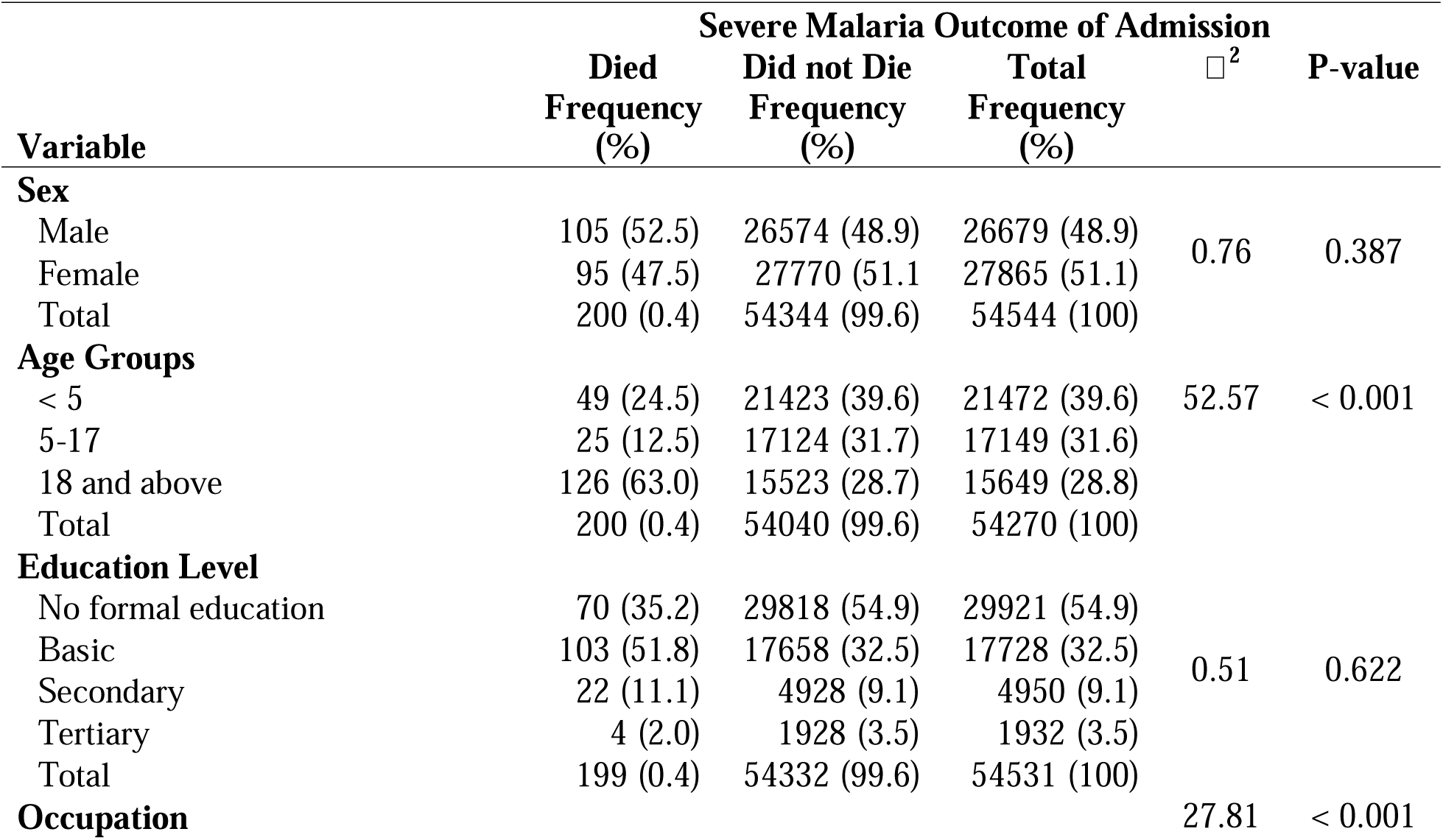
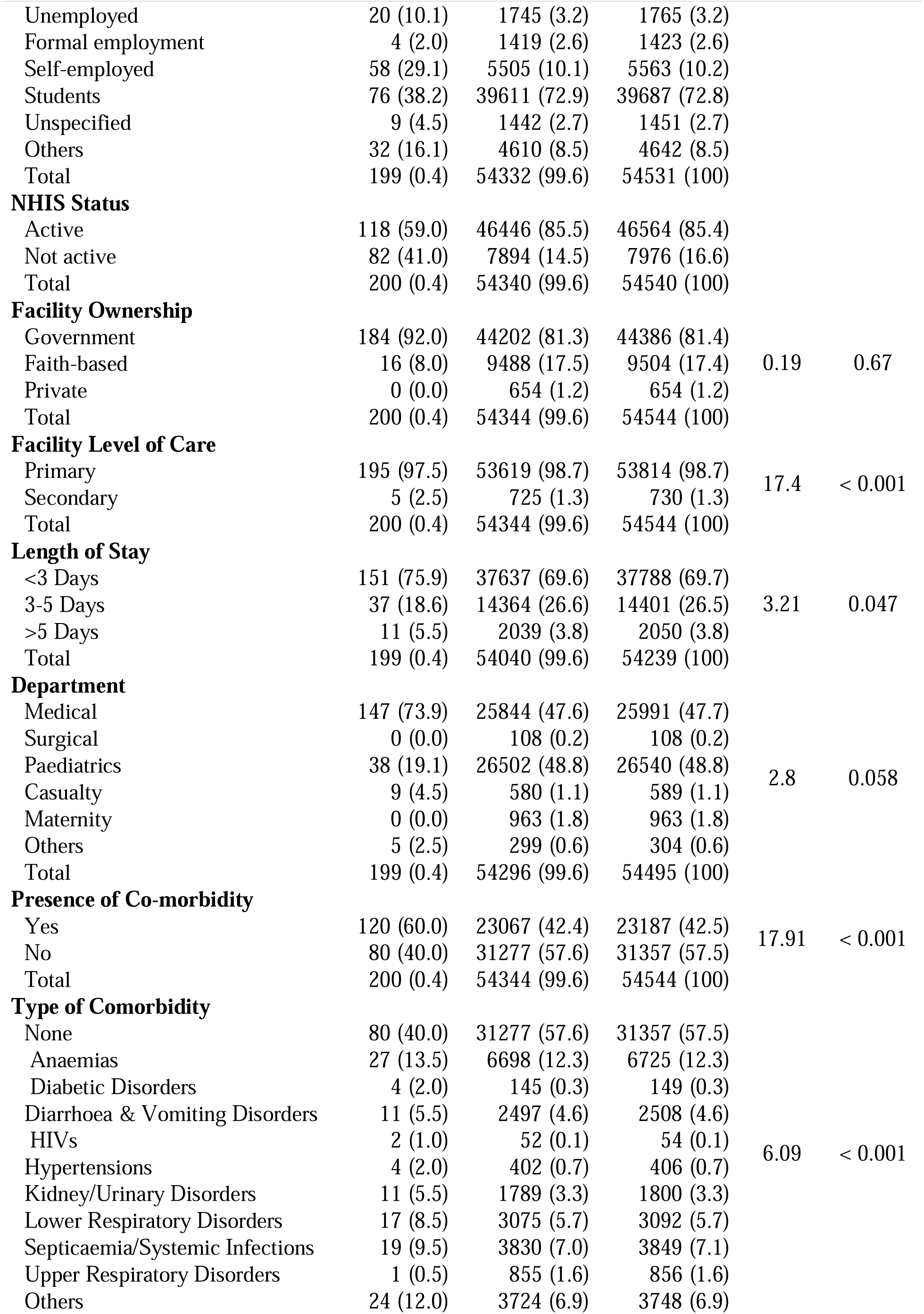
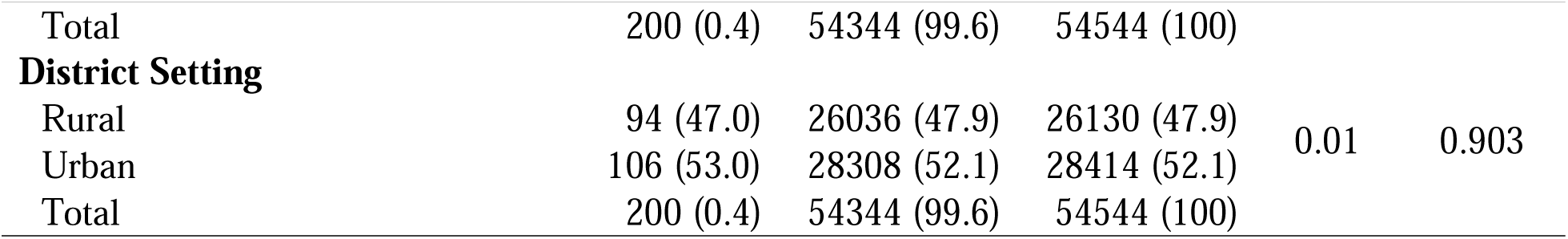
Bivariate analysis of patient characteristics and severe malaria admission outcomes in the Ashanti region, Ghana, 2018-2022.

After statistically significant and theoretically important variables were entered into a multivariable model using Firth’s penalised likelihood logistic regression, given that the prevalence of severe malaria deaths is 0.4% (less than 5%). Upon holding other variables constant, sex, age group, occupations, NHIS status, facility ownership, length of stay, departments, and type of comorbidities were significantly associated with the outcome. Males were 1.4 times more at odds of death compared to females (aOR = 1.39, 95% CI: 1.04 – 1.86, p = 0.026). Compared to the under-5-year-olds, the 5-17 age group exhibited 44% lower odds of death (aOR = 0.56, 95% CI: 0.33 – 0.94, p = 0.029). In comparison to the unemployed, students had 70% lower odds of death (aOR = 0.30, 95% CI: 0.16 – 0.58, p < 0.001). Patients with active NHIS status had 66% lower odds of death compared to those without active status (aOR = 0.33, 95% CI: 0.25 – 0.45, p < 0.001). Patients treated in faith-based facilities had 62% lower odds of death compared to those treated in government facilities (aOR = 0.38, 95% CI: 0.23 – 0.65, p < 0.001). Patients admitted for 3-5 days had 33% lower odds of death compared to those on admission for less than 3 days (aOR = 0.67, 95% CI: 0.47 – 0.97, p = 0.033). Compared to the paediatric department, the medical department showed 2 times higher odds of death (aOR = 0.2.38, 95% CI: 1.48 – 3.84, p < 0.001), while those in the casualty department had more than 5 times higher odds of dying (aOR = 5.46, 95%: 2.48 – 11.99, p < 0.001). Compared to inpatients without comorbidities, those with comorbidities had 2.3 times higher odds of death (aOR = 2.75, 95% CI: 1.77 – 4.50, p < 0.001) (Table 3). Assessment of multicollinearity using variance inflation factors (VIFs) indicated generally independent predictors (mean VIF = 1.88; all variables below the threshold of 10). Model diagnostics indicated good fit (AIC = 2295.48, BIC = 2517.86), and ROC analysis demonstrated acceptable discrimination (AUC = 0.81, 95% CI: 0.78 – 0.84) (Table 3).

**Table 3.** Penalized multivariable logistic regression analysis for the association between severe malaria admission outcomes and inpatient characteristics in the Ashanti region, 2018 – 2022.

| Variable | Crude OR (95% CI) | P-Value | Adjusted OR (95% CI) | P-Value |
| --- | --- | --- | --- | --- |
| **Sex** |  |  |  |  |
| Male | 1.15 (0.87, 1.52) | 0.310 | 1.39 (1.04, 1.86) | 0.026 |
| Female | 1 |  | 1 |  |
| **Age Groups** |  |  |  |  |
| < 5 | 1 |  | 1 |  |
| 5-17 | 0.64 (0.40, 1.04) | 0.072 | 0.56 (0.33, 0.94) | 0.029 |
| 18 and above | 3.53 (2.54, 4.90) | < 0.001 | 1.31 (0.70, 2.44) | 0.399 |
| **Education Level** |  |  |  |  |
| Basic | 1 |  | 1 |  |
| Secondary | 1.14 (0.71, 1.84) | 0.581 | 0.95 (0.57, 1.59) | 0.846 |
| Tertiary | 0.58 (0.23, 1.51) | 0.270 | 0.44 (0.15, 1.31) | 0.140 |
| No formal education | 0.87 (0.64, 1.18) | 0.366 | 1.36 (0.97, 1.90) | 0.076 |
| **Occupation** |  |  |  |  |
| Unemployed | 1 |  | 1 |  |
| Formal employment | 0.27 (0.10, 0.75) | 0.012 | 0.41 (0.13, 1.27) | 0.123 |
| Self-employed | 0.90 (0.55, 1.50) | 0.698 | 0.70 (0.41, 1.17) | 0.174 |
| Students | 0.16 (0.10, 0.26) | < 0.001 | 0.30 (0.16, 0.58) | < 0.001 |
| No Information/Unspecified | 0.56 (0.26, 1.21) | 0.142 | 0.48 (0.22, 1.04) | 0.064 |
| Others | 0.60 (0.34, 1.05) | 0.072 | 0.55 (0.31, 0.96) | 0.035 |
| **NHIS Status** |  |  |  |  |
| Active | 0.24 (0.18, 0.32) | < 0.001 | 0.33 (0.25, 0.45) | < 0.001 |
| Not active | 1 |  | 1 |  |
| **Facility Ownership** |  |  |  |  |
| Government | 1 |  | 1 |  |
| Faith-based | 0.42 (0.25, 0.69) | 0.001 | 0.38 (0.23, 0.65) | < 0.001 |
| Private | 0.18 (0.01, 2.94) | 0.231 | 0.25 (0.02, 3.97) | 0.323 |
| **Facility Level of Care** |  |  |  |  |
| Primary | 1 |  | 1 |  |
| Secondary | 2.08 (0.89, 4.87) | 0.092 | 2.07 (0.79, 5.43) | 0.137 |
| **Length of Stay** |  |  |  |  |
| <3 Days | 1 |  | 1 |  |
| 3-5 Days | 0.65 (0.45, 0.93) | 0.018 | 0.67 (0.47, 0.97) | 0.033 |
| >5 Days | 1.40 (0.77, 2.55) | 0.272 | 1.26 (0.68, 2.32) | 0.458 |
| **Department** |  |  |  |  |
| Paediatrics | 1 |  | 1 |  |
| Maternity | 0.36 (0.02, 5.82) | 0.470 | 0.26 (0.02, 4.32) | 0.346 |
| Surgical | 3.17 (0.19, 51.96) | 0.418 | 1.77 (0.10, 30.14) | 0.695 |
| Medical | 3.93 (2.75, 5.60) | < 0.001 | 2.38 (1.48, 3.84) | < 0.001 |
| Casualty | 11.27 (5.51, 23.02) | < 0.001 | 5.46 (2.48, 11.99) | < 0.001 |
| Others | 12.64 (5.14, 31.11) | < 0.001 | 7.51 (0.02, 19.49) | < 0.001 |
| **Presence of Co-morbidity** |  |  |  |  |
| Yes | 2.03 (1.52, 2.69) | < 0.001 | 2.30 (1.72, 3.08) | < 0.001 |
| No | 1 |  | 1 |  |
| **District Setting** |  |  |  |  |
| Rural | 1 |  | 1 |  |
| Urban | 1.04 (0.78, 1.37) | 0.800 | 0.83 (0.63, 1.11) | 0.220 |
| **Model Diagnostics** |  |  |  |  |
| VIF |  |  | 2.37 (1.01 – 9.73)<sup>†</sup> |  |
| ROC/AUC |  |  | 0.81 (0.78, 0.84)<sup>‡</sup> |  |
| AIC |  |  | 2295.48<sup>1</sup> |  |
| BIC |  |  | 2517.86<sup>2</sup> |  |

Notes: 1 – Reference; OR – Odds Ratio; aOR – Adjusted Odds Ratio; 95% CI – 95% Confidence Interval; VIF - Variance Inflation Factor; ROC/AUC – Receiver Operating Characteristic curve/ Area Under the Curve; AIC – Akaike Information Criteria; BIC – Bayesian Information Criteria.
<sup>†</sup> Mean (range) – mean VIF = 2.37, and range of VIF all less than the threshold of 10 indicates acceptable collinearity.
<sup>‡</sup> AUC (95% CI) – AUC = 0.81 suggest a strong model performance.
<sup>1</sup> AIC = 2295.48 supports an adequate fit of the model.
<sup>2</sup> BIC = 2517.86 supports an adequate fit of the model.

Sensitivity analysis with the survey-adjusted logistic regression showed consistent direction and significance across covariates except for sex (aOR = 1.39, 95% CI: 0.97 – 2.00, p = 0.071) and length of stay (aOR = 0.67, 95% CI: 0.43 – 1.03, p = 0.070), where significance changed (See Additional file 1). Postestimation for the survey-adjusted logistic regression model, using the Adjusted Wald test to assess the joint significance of predictors, indicated a significant influence on the outcome variable (F (_21, 29_) = 28.41, P-value < 0.001). The link test showed significant predicted values (2.82, 95% CI: 1.13 – 7.05, p = 0.028), which was in contrast to the square predicted value (1.00, 95% CI: 0.92 – 1.10, p = 0.935), and ROC analysis also demonstrated good discrimination (AUC = 0.80, 95% CI: 0.77 – 0.84) (See Additional file 1).

## Discussions

This study aimed to describe the characteristics of inpatient severe malaria and identify factors associated with admission outcomes using secondary data from DHIMS 2 from 2018 to 2022. The study observed an overall case fatality rate of 0.4% during the period under study. Several factors were found to be associated with severe malaria mortality, including sex, age distribution, occupation, NHIS status, facility ownership, length of hospital stay, admitting department, and the presence of comorbid conditions. While education, facility level of care, and district setting were relevant characteristics, they did not emerge as significant predictors of inpatient outcomes in this analysis.

The study observed a higher proportion of female admissions for severe malaria, consistent with findings from Rwanda and Ghana, where females accounted for 59% and 62% of malaria admissions, respectively (41,42). This pattern may reflect differences in health-seeking behaviour, with women more likely to pursue medical attention (43). In addition, women may have increased exposure to malaria vectors during peak transmission hours (42). The significant association observed between sex and mortality suggests that biological, behavioural, or care-seeking differences may be related to inpatient outcomes in the region. However, causal mechanisms cannot be determined from this study.

Age was significantly associated with inpatient mortality, with children under five years accounting for about 40% of severe malaria admissions and approximately 25% of deaths, aligning with other studies (44,45). This finding is consistent with the known vulnerability of young children, who have underdeveloped immune systems and declining maternal immunity (46). Older children aged 5–17 years had lower odds of mortality compared to those under five, consistent with other literature (47). This pattern may reflect partial immunity development, stronger physiological resilience, and improved ability to report symptoms, which may facilitate earlier presentation and care (48–50).

Although fewer than half of severe malaria admissions involved patients with comorbid conditions, comorbidities were strongly associated with higher mortality. Patients with comorbidities had more than twofold higher odds of death compared to those without comorbid conditions, consistent with existing literature (5,51–53). Conditions such as diabetes, septicaemia, and other systemic infections have been widely reported to be associated with more severe disease and poorer outcomes (5,54–56). These findings support current recommendations that emphasize the importance of screening for and managing co-infections and other comorbidities in patients admitted with severe malaria (5)

Socioeconomic factors, particularly occupation and NHIS status, were associated with inpatient severe malaria outcomes. Occupation remained significant in multivariable analysis, with students showing lower mortality odds compared to those unemployed. Unlike a study in Liberia that found no association between occupation and malaria severity (57), our results suggest occupation influences inpatient outcomes in Ghana, likely through differences in health-seeking behavior and financial access to care. This finding aligns with patterns observed in similar settings, where employment status influences health-seeking behavior, financial access to care, and awareness of early symptoms (58–60). Unemployed individuals often face greater barriers, including limited resources and delayed presentation, which may contribute to poorer outcomes (61). These associations likely reflect differences in socioeconomic stability and healthcare access rather than occupation as a direct cause of mortality (10,62–64). Active NHIS membership was associated with substantially lower odds of mortality, consistent with other studies (37). Health insurance is likely associated with earlier presentation, reduced financial barriers, and improved continuity of care (21,65,66). However, this study cannot establish that NHIS enrollment directly reduces mortality, and residual confounding by socioeconomic status and disease severity at presentation may partly explain this association.

Patterns in facility ownership were also associated with differences in inpatient mortality. Most severe malaria cases were managed in government facilities, reflecting the central role of the public health system (7,67). Patients admitted to faith-based facilities had lower odds of mortality compared to those in government facilities, consistent with findings from other studies (37). However, this association should be interpreted with caution. It may reflect differences in case mix, referral patterns, patient volume, or severity of illness at admission, rather than differences in quality of care alone (68) Further assessment is needed to understand whether these observed differences are driven by facility practices, patient characteristics, or health system factors. Nonetheless, identifying high-performing facilities and understanding their operational practices may provide useful insights for quality improvement across facilities, for example, through benchmarking frameworks such as the SafeCare Quality Improvement Standards (69).

The wards of admission were also associated with inpatient mortality. Patients admitted to medical and casualty departments had higher odds of death compared to those admitted to paediatric departments. This pattern is consistent with findings from Uganda and other settings (70) and may reflect higher disease severity, delayed presentation, or more complex comorbidities among patients triaged to medical and emergency units (5) These findings may not necessarily imply differences in quality of care across departments, but rather as indicators of differences in patient profiles and illness severity at admission.

Length of hospital stay was also associated with mortality. Most patients were hospitalized for less than three days. Patients admitted for 3–5 days had lower odds of mortality compared to those with shorter stays. However, this finding should be interpreted cautiously, as it may reflect reverse causality. Patients who died early would, by definition, have shorter lengths of stay. Therefore, shorter admission duration may be a marker of early death rather than premature discharge. As such, this association does not necessarily imply that longer admission directly improves survival, but it does highlight the importance of close monitoring during the early phase of admission for severe malaria (71–75).

The combined use of Firth’s penalized likelihood logistic regression and survey-adjusted logistic regression as a sensitivity analysis strengthens confidence in the robustness of the observed associations (40). Firth’s method helped address bias related to the very low prevalence of inpatient deaths, while the survey-adjusted models accounted for clustering at the facility level (76–78). The consistency in the direction and magnitude of most estimates across models supports the stability of the findings, even where statistical significance differed slightly for some variables.

Despite the large dataset, several important limitations should be considered. First, the cross-sectional design does not allow for assessment of temporal relationships, and causal inferences cannot be made. Second, DHIMS 2 does not capture key clinical severity indicators, such as parasite density, level of consciousness, haemoglobin levels, organ dysfunction, or time to treatment. As a result, residual confounding by disease severity at presentation is likely and may partly explain associations related to facility type, department of admission, and length of stay. Finally, exclusion of Komfo Anokye Teaching Hospital, a major referral centre, may limit generalizability to the broader Ashanti Region. These limitations highlight the need for cautious interpretation of the findings and underscore the importance of complementary prospective and clinical audit studies to better understand causal pathways.

### Conclusion

This study described the characteristics of inpatients with severe malaria and identified factors associated with mortality following admission in the Ashanti Region of Ghana, using routine surveillance data from DHIMS 2 for the period spanning 2018 to 2022. The study demonstrated significant associations between mortality and sex, age distribution, occupation, NHIS status, facility ownership, as well as admitting department, duration of stay in the health facility, and the presence of comorbid conditions. Active NHIS status, age between 5 and 17 years, admission to a faith-based facility, and hospital stays of 3 to 5 days were associated with lower odds of mortality. Meanwhile, male sex, unemployment, admission to medical and casualty departments, and the presence of comorbidities were associated with higher odds of death among patients admitted with severe malaria.

These findings imply that training healthcare providers to recognize demographic-specific vulnerabilities, such as the heightened risk among older patients and those with comorbid conditions, could further enhance patient outcomes through improved triage and early clinical intervention. In addition, facilities with a high proportion of older or clinically complex severe malaria admissions should be identified and resourced in terms of training, diagnostics, medications, and other essential supplies to improve outcomes. The observed protective association of NHIS enrollment emphasizes the need for targeted policies to enhance health insurance accessibility and coverage, particularly for low-income populations. In this vein, the Government of Ghana’s plan to make primary healthcare accessible to all is likely to influence outcomes among patients admitted for severe malaria; however, these efforts may prove counterproductive if they are not appropriately managed and resources are not properly targeted to facilities and populations with the greatest need.

Given the strong association between comorbidities and severe malaria admission outcomes, there is a need for severe malaria guidelines to include routine screening for common comorbid conditions and integrated clinical management. In addition, standardizing severe malaria treatment protocols using best practices observed in paediatric departments, and benchmarking quality strategies used by faith-based facilities, may support improvements in inpatient outcomes across facility types.

Future research should examine the long-term impact of NHIS enrollment on malaria-related mortality using longitudinal designs to better assess temporal relationships. Investigations into disparities in care processes and patient case mix across different facility types could provide valuable insights into variations in patient outcomes. Further studies assessing the influence of non-communicable diseases and other chronic conditions on malaria prognosis may help refine treatment strategies for at-risk populations.

## Supporting information

Supplemental Sensitivity analysis and diagnostics

## Declarations

### Ethics approval and consent to participate

The study received ethical approval from the Ghana Health Service Ethics Review Committee with approval number GHS-ERC:044/06/24 obtained on 5^th^ July 2024. The Ethics Committee waived the need for written informed consent from each participant due to the use of secondary data, which were de-identified and had minimal risk. Further permission to use the data was obtained from the head of the Centre for Health Information Management, Ghana (CHIM) and the Ashanti Regional Health Directorate. This study was conducted in accordance with the Declaration of Helsinki.

### Consent for publication

Not applicable

### Availability of data and materials

The datasets used and/or analysed during the current study are available from the corresponding author on reasonable request.

## Authors’ contributions

HAB and CEY conceptualised and designed the study. MMO and CEY analysed the data with support from HAB. CEY and MMO drafted the initial manuscript. HAB and BS critically reviewed and revised the manuscript for important intellectual content. All authors read and approved of the final manuscript.

## Acknowledgements

We are grateful to the Ghana Health Service and Centre for Health Information Management, Ghana, for granting access to the data.

## Additional files

Additional file 1: Survey-adjusted multivariable logistic regression analysis for the association between severe malaria admission outcomes and inpatient characteristics in the Ashanti region, 2018 – 2022.

