## Supplemental Sensitivity analysis and diagnostics for "Factors Associated with Outcomes of Inpatient Severe Malaria Cases in the Ashanti Region, Ghana: An Analytic Cross-sectional Study using Routine Surveillance Data, 2018 to 2022"

**Supplementary Materials**

**Table 1.** **Variable Table**

| **Variable name** | **Definition** | **Type of Variable** | **Categories** |
| --- | --- | --- | --- |
| **Sex** | Biological sex | Categorical | Female  Male |
| **Age** | Age of the patient (in years) | Categorical | < 5  5-17  18 and above |
| **Educational level** | The level of education of the patient (grouped into different categories) | Categorical | No formal education  Basic  Secondary  Tertiary |
| **Occupation** | The kind of work the patient does (recategorized into groups merging occupations with similar characteristics) | Categorical | Unemployed  Formal Employment  Self-employed  Student  Unspecified  Others |
| **NHIS Status** | Patient’s active membership on the NHIS at the time of admission | Categorical | Active  Not Active |
| **Co-morbidities** | Presence of any other existing disease at the time or during the severe malaria. Categories adapted and modified from the NHIS G-DRG coding system (ICD10/ICD11) | Categorical | None  Anaemias  Diabetic Disorders  Diarrhoea & Vomiting Disorders  HIVs  Hypertension  Kidney/Urinary Disorders  Lower Respiratory Disorders  Septicaemia/Systemic Infections  Upper Respiratory Disorders  Others |
| **Facility Ownership** | Type of ownership of the facility | Categorical | Government  Quasi-governmental  Mission  Private |
| **Level of facility** | Level of operation of the facility according to NHIA/GHS criteria | Categorical | Primary Hospital  Secondary Hospital  Tertiary Hospital |
| **Department** | Unit or department in the hospital where the admission took place | Categorical | Medical  Surgical  Pediatric  Casualty  Maternity  Others |
| **Length of stay** | How long the patient stays on admission during an episode of severe malaria (in days) | Categorical | < 3  3-5  >5 |
| **District Setting** | Classifications of District Setting | Categorical | Urban  Rural |

**Table 2. Grouping of Comorbities using G-DRG.**

| **Comorbidities** | **Core G-DRG** | **ICD-10 Code** | **Details of Diagnoses/Indications for procedure(s)** |
| --- | --- | --- | --- |
| Anaemias/Anaemic Disorders | MEDI06 | D50.9 | Iron deficiency anaemia |
|  | PAED06 | D55.0 | Anaemia (G6PD) |
|  |  | D57 | Sickle Cell Disorders |
|  |  | D57.0 | Sickle Cell Crises |
|  |  | D57.1 | Sickle Cell Crisis |
|  |  | D57.1 | Haemoglubinopathy with Crisis |
|  |  | D57.1 | Sickle Cell |
|  |  | D57.1 | Sickle Cell Anaemia |
|  |  | D57.1 | Sickle Cell Disease |
|  |  | D58.9 | Anaemia Heamolytic Hereditary |
|  |  | D58.9 | Hereditary Haemolytic Anaemia, unspecified |
|  |  | D59.9 | Acquire Haemolytic Anaemia, Unspecified |
|  |  | D59.9 | Anaemia Haemolytic Acquired |
|  |  | D61.9 | Anaemia Aplastic |
|  |  | D64.9 | Anaemia |
|  |  | O99.0 | Anaemia in Pregnancy |
|  |  |  | Sickle Cell Anaemia without Crises |
|  |  | O99.0 | Sickle Cell Disease in Pregnancy |
| Hypertensions | MEDI32 | I10 | Hypertension |
|  | PAED40 | I11.9 | Hypertensive Heart Disease |
|  | OBGY09 | O13 | Gestational Hypertension |
|  |  | O13 | Hypertension Gestationed |
|  |  | O13 | Pregnancy-induced Hypertension |
|  |  | I67 | Encephalopathy Hypertensive |
| HIVs | MEDI38 | B23 | HIV Infection |
|  | PAED45 | B24 | HIV/AIDS |
| Diabetic Disorders | MEDI02 | E14 | Diabetes |
|  | MEDI03 | E14 | Diabetes Mellitus |
|  | PAED02 | E14.0 | Diabetic Ketoacidosis |
|  | PAED03 | E14.2 | Diabetic Nephropathy |
|  |  | E14.0 | Hypoglycaemic Coma |
|  |  | E14.5 | Diabetic Foot |
|  |  | E87.0 | Hyperosmolar Nonketotic Hyperglycaemia (HONK) |
|  |  | R73.9 | Hyperglycaemia |
| Renal/Kidney/Urinary Tract Disorders | MEDI19 | N00 | Nephritis (Acute) |
|  | MEDI20 | N04 | Nephrotic Syndrome |
|  | MEDI21 | N05 | Nephritic Syndrome |
|  |  | N05 | Nephrotic Syndrome |
|  |  | N10 | Pyelonephritis (Acute) |
|  |  | N12 | Pyelonephritis |
|  |  | N12 | Pyelonephritis (Unspecified) |
|  |  | N13.0 | Hydronephrosis with Ureteropelvic Junction Obstructions |
|  |  | N13.9 | Obstruction Urinary |
|  |  | N13.9 | Chronic Kidney Disease |
|  |  | N19 | Renal Failure |
|  |  | N28.9 | Disease of Kidney (Unspecified) |
|  |  | N32.8 | Cystocele Male |
|  |  | N34.0 | Urethral Abscess |
|  |  | N39.0 | Infection of Urinary Tract (UTI) |
|  |  | N39.9 | Disease of the Urinary System (Unspecified) |
|  |  | N40 | Benign Prostrate Hyperplasia |
|  |  | O86.2 | Genitourinary Tract Infection in Pregnancy |
|  |  | Q61.4 | Renal Dysplesia |
|  |  | R33 | Retention of Urine |
| Septicaemia/Systemic Infections | MEDI30 | A01.0 | Enteric Fever |
|  | PAED38 | A01.0 | Typhoid Fever |
|  | PAED14 | A01.0 | Typhoid Perforation |
|  |  | A02.1 | Salmella Sepsis |
|  |  | A33.0 | Neonatal Tetanus |
|  |  | A39.0 | Cerebrospinal Meningitis (CSM) |
|  |  | A39.0 | Meningitis Cerebrospinal (CSM) |
|  |  | A41.9 | Sepsis Unspecified |
|  |  | A41.9 | Septicaemic Shock |
|  |  | A95.9 | Yellow Fever |
|  |  | G00.1 | Pneumococcal Meningitis |
|  |  | G00.9 | Bacterial Meningitis |
|  |  | G04.9 | Encephalitis |
|  |  | G04.9 | Encephalitis (Unspecified) |
|  |  | M84.9 | Osteomyelitis |
|  |  | O85 | Child Birth Sepsis |
|  |  | O85 | Postpartum Sepsis |
|  |  | O85 | Pueperial Sepsis |
|  |  | P36.9 | Cord Sepsis Neonatal |
|  |  | P36.9 | Sepsis of Cord in Newborn |
|  |  | P36.9 | Neonatal Sepsis |
|  |  | M00.9 | Septic Arthritis |
|  |  | O07.5 | Septic Abortion |
|  |  | R57.8 | Septic Shock |
| Diarrhoeal & Vomiting Disorders | MEDI23 | A00 | Cholera |
|  | PAED31 | A03.9 | Bacillary Dysentery |
|  |  | A03.9 | Dysentery Bacilliary |
|  |  | A03.9 | Shigellosis |
|  |  | A06.0 | Dysentery Amoebic (Acute) |
|  |  | A06.9 | Amoebiasis |
|  |  | A09 | Diarrhoea |
|  |  | A09 | Dysentery |
|  |  | A09 | Enema Colitis |
|  |  | A09 | Enteritis |
|  |  | A09 | Gastroenteritis |
|  |  | K52.2 | Gastroenteritis and Colitis (Allergic) |
| Lower Respiratory Disorders | MEDI22 | J15.7 | Atypical Pneumonia |
|  | MEDI31 | J18.0 | Bronchopneumonia |
|  | PAED12 | J18.1 | Pneumonia Lobar |
|  | PAED30 | J18.9 | Mild - Moderate Pneumonia |
|  | PAED39 | J18.9 | Pneumonia |
|  |  | J18.9 | Severe Pneumonia |
|  |  | J20.9 | Bronchitis (Acute) |
|  |  | J21.9 | Bronchiolitis |
|  |  | J22 | Infection Lower Respiratory Tract (Acute) |
|  |  | J40. | Bronchitis |
|  |  | J40 | Brochitis (Unspecified) |
|  |  | J42 | Infection of Lower Respiratory Tract (Chronic) |
|  |  | J45.9 | Asthma |
|  |  | J45.9 | Bronchial Asthma |
|  |  | J69.0 | Aspiration Pneumonia |
|  |  | J81 | Pulmonary Oedema |
|  |  | J90 | Pleural Effusion |
|  |  | J98.0 | Bronchospasm |
|  |  | J98.8 | Acute Chest Infection |
|  |  | J98.8 | Infection of Respiratory Tract (RTI) |
|  |  | O99.5 | Bronchopneumonia in pregnancy |
|  |  | P21.9 | Birth Asphyxia |
|  |  | J98.9 | Respiratory Disoder, Unspecified |
|  |  | A16.2 | Tuberculosis Pulmonary |
|  |  | A16.9 | Kochs |
|  |  | A16.2 | Tuberculosis |
|  |  | A37.9 | Pertussis |
|  |  | O99.5 | Chest Infection in Pregnancy |
|  |  | R05 | Cough |
|  |  | R06.8 | Respiratory Insufficiency |
|  |  | P22.0 | Respiratory Distress Sydrome |
|  |  | O99.5 | Cyesis with Asthmatic Attack |
| Upper Respiratory Disorders |  | J02.9 | Pharyngitis (Acute) |
|  |  | J03.9 | Acute Tonsilitis (Unspecified) |
|  |  | J03.9 | Acute Tonsilitis |
|  |  | J06.9 | Infection of Upper Respiratory Tract |
|  |  | J06.9 | Infection of Upper Respiratory Tract (Acute) |
|  |  | J06.9 | Upper Respiratory Tract Infection |
|  |  | J11.1 | Influeza |
|  |  | J32.9 | Chronic Sinusitis Unspecified |
|  |  | J32.9 | Rhinosinusitis |
|  |  | J32.9 | Sinusitis |
|  |  | J34.8 | Disease of the Nose |
|  |  | J34.7 | Disease of Larynx |
|  |  | J39.2 | Disease of Pharynx |
|  |  | J39.8 | Infection of Upper Respiratory (Chronic) |
|  |  | K14.0 | Glossitis |
|  |  | R07.0 | Painful Throat |

**Table 3. Survey-adjusted multivariable logistic regression analysis for the association between severe malaria admission outcomes and inpatient characteristics in the Ashanti region, 2018 – 2022.**

| **Variable** | **Crude OR (95% CI)** | **P-Value** | **Adjusted OR (95% CI)** | **P-Value** |
| --- | --- | --- | --- | --- |
| **Sex** |  |  |  |  |
| Male | 1.16 (0.83, 1.61) | 0.388 | 1.39 (0.97, 2.00) | 0.071 |
| Female | 1 |  | 1 |  |
| **Age Groups** |  |  |  |  |
| < 5 | 1 |  | 1 |  |
| 5-17 | 0.64 (0.41, 1.00) | 0.052 | 0.55 (0.33, 0.91) | 0.022 |
| 18 and above | 3.55 (2.37, 5.30) | < 0.001 | 1.32 (0.67, 2.63) | 0.418 |
| **Education Level** |  |  |  |  |
| Basic | 1 |  | 1 |  |
| Secondary | 1.12 (0.58, 2.17) | 0.719 | 0.93 (0.49, 1.78) | 0.826 |
| Tertiary | 0.52 (0.20, 1.34) | 0.174 | 0.40 (0.14, 1.16) | 0.090 |
| No formal education | 0.87 (0.50, 1.53) | 0.625 | 1.36 (0.86, 2.14) | 0.181 |
| **Occupation** |  |  |  |  |
| Unemployed | 1 |  | 1 |  |
| Formal employment | 0.25 (0.09, 0.70) | 0.010 | 0.38 (0.13, 1.13) | 0.080 |
| Self-employed | 0.92 (0.58, 1.46) | 0.719 | 0.71 (0.44, 1.17) | 0.173 |
| Students | 0.17 (0.10, 0.27) | < 0.001 | 0.31 (0.16, 0.59) | 0.001 |
| No Information/Unspecified | 0.54 (0.24, 1.23) | 0.142 | 0.46 (0.22, 0.95) | 0.037 |
| Others | 0.61 (0.34, 1.09) | 0.093 | 0.55 (0.28, 1.07) | 0.077 |
| **NHIS Status** |  |  |  |  |
| Active | 0.24 (0.16, 0.38) | < 0.001 | 0.33 (0.22, 0.51) | < 0.001 |
| Not active | 1 |  | 1 |  |
| **Facility Ownership** |  |  |  |  |
| Government | 1 |  | 1 |  |
| Faith-based | 0.41 (0.23, 0.71) | 0.002 | 0.37 (0.20, 0.69) | 0.002 |
| Private | - |  | - |  |
| **Facility Level of Care** |  |  |  |  |
| Primary | 1 |  | 1 |  |
| Secondary | 1.90 (1.39, 2.59) | < 0.001 | 1.86 (0.77, 4.49) | 0.161 |
| **Length of Stay** |  |  |  |  |
| <3 Days | 1 |  | 1 |  |
| 3-5 Days | 0.64 (0.41, 0.99) | 0.048 | 0.67 (0.43, 1.03) | 0.070 |
| >5 Days | 1.34 (0.75, 2.42) | 0.319 | 1.21 (0.65, 2.26) | 0.538 |
| **Department** |  |  |  |  |
| Paediatrics | 1 |  | 1 |  |
| Maternity | - |  | - |  |
| Surgical | - |  | - |  |
| Medical | 3.97 (2.53, 6.21) | < 0.001 | 2.38 (1.19, 4.76) | 0.015 |
| Casualty | 10.82 (3.56, 32.94) | < 0.001 | 5.21 (1.76, 15.41) | 0.004 |
| Others | 11.66 (3.91, 34.81) | < 0.001 | 6.91 (2.57, 18.53) | < 0.001 |
| **Presence of Co-morbidity** |  |  |  |  |
| Yes | 2.03 (1.44, 2.86) | < 0.001 | 2.31 (1.73, 3.08) | < 0.001 |
| No | 1 |  | 1 |  |
| **District Setting** |  |  |  |  |
| Rural | 1 |  | 1 |  |
| Urban | 1.04 (0.57, 1.88) | 0.903 | 0.84 (0.51, 1.36) | 0.460 |
| **Postestimations/Diagnostics** |  |  |  |  |
| Adjusted Wald Test |  |  | *F_(21, 29)_ = 28.41* | *< 0.001* |
| ROC/AUC |  |  | *0.80 (0.77, 0.84)* |  |
| **Link Test** |  |  |  |  |
| Predicted |  |  | *2.82 (1.13, 7.05)* | *0.028* |
| Square predicted |  |  | *1.00 (0.92, 1.10)* | *0.935* |

*Note: Supplementary Table 3. Sensitivity analysis using survey‑adjusted logistic regression. Candidate variables identified through descriptive and bivariate analyses were re-entered into a multivariable survey‑adjusted logistic regression model to account for clustering and complex design effects. Findings were consistent in direction and magnitude with the main Firth penalised likelihood model, confirming robustness. The two instances of difference, sex and length of stay, only showed slight changes in significance, which is regarded as no meaningful change as discussed in the main text. This table is provided in the supplementary materials to maintain the conciseness of the main text while ensuring transparency of the sensitivity analyses.*

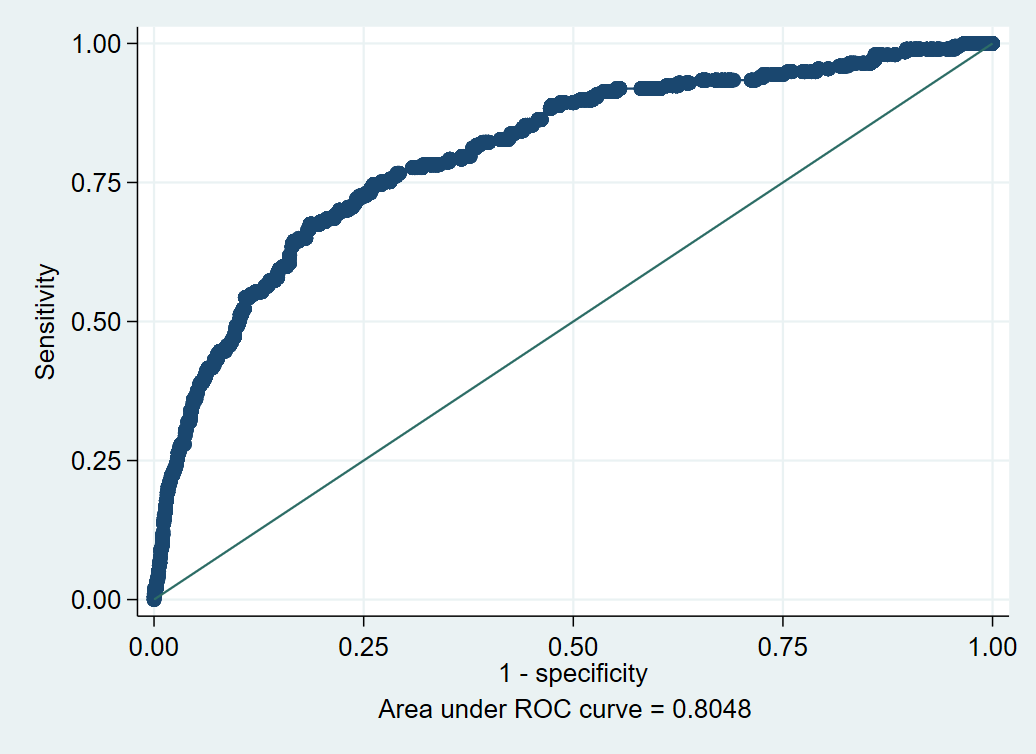

Figure 1. ROC curve for the survey-adjusted multivariable logistic regression model for predictors of severe malaria admission outcomes.

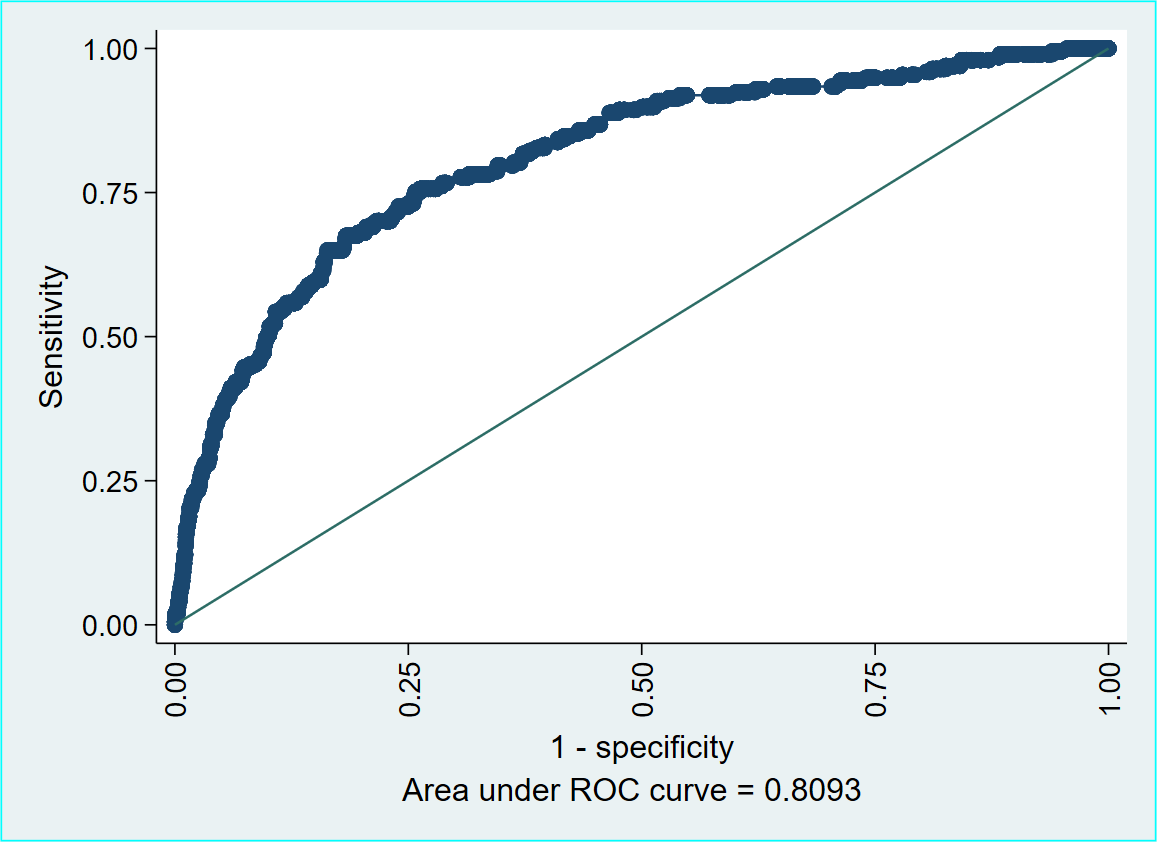

Figure 2. ROC curve for the multivariable Firth's penalized likelihood logistic regression model for predictors of severe malaria admission outcomes.

.
